# Seroprevalence of Varicella Zoster virus in China: an age-stratifed systematic review and meta-analysis

**DOI:** 10.1101/19009449

**Authors:** Li Wang, Fu Sun, Siyang Liu, Hongyi Zhang, Bo Lu, Xiaoli Shao, Minxin Zhang, Zhengwei Li

## Abstract

**OBJECTIVES:** the aim of this meta-analysis was pooled on the comprehensive information of the epidemiology of VZV infection and analyzed the seropositivity of VZV-IgG antibodies in different age groups in China, so as to arouse people’s attention on VZV.

**DESIGN:** Systematic review

**DATA SOURCES:** Sources included on PubMed, the China National Knowledge Infrastructure (CNKI) database, WANFANG database and the Chinese Scientific Journals Full-Text Database (CQVIP) from 1997 to 2019.

**ELIGIBILITY CRITERIA:** (1) the Chinese and English literatures of study on varicella-zoster virus immunization epidemics in different regions of China; (2) The literature has obvious age stratification; (3) The subjects are general people; (4) VZV sero-prevalence in the population investigated by the literature research institute. Exclusion criteria: Any study that did not contain these information was excluded.

**DATA EXTRACTION AND SYNTHESIS:** Two evaluators independently retrieved documents and extracted data. Information about the study design, eligible population, age and gender distribution, inclusion and exclusion criteria were checked. When they disagreed, they could solve the problem by discussion or by soliciting opinions from third parties.

**RESULTS:** Literatures were screened according to the inclusion criteria and 10 studies from 1997 to 2019 with a total of 11666 individuals were included. The overall VZV seroprevalence in the Chinese population was 65.80% (95% CI; 56.5% - 75.1%), and the peak prevalence was seen in the age 36-45 93.50% (95% CI; 0.917-0.953) while the VZV seroprevalence rate (82.20%, 95% CI: 0.544-1.099) was not increased in individuals of 45 and older.

**CONCLUSION:** The incidence of VZV increases with age, and there was no significant difference between different genders or regions. This results can provide epidemiological evidence for the prevention and treatment of VZV.

**Strengths and limitations of this study:**

- A broad search strategy and systematic review methodology were used to generate this comprehensive review on advice for prevention and treatment of VZV.
- Every inclusion article was assessed with a risk of bias tool and/or a quality assessment tool.
- Because of relatively small sample size, the incomplete data, and the different research objects, there are some bias in the outcomes, which limits the ability to make discrete conclusions.
- There were not enough articles identified to perform a meta-analysis.

## Introduction

Varicella-Zoster Virus (VZV) is the pathogen of human varicella and herpes zoster, and human is the only host(**Davison, 1986)**. An average of 4.2 million severe varicella cases cause hospitalization or death worldwide each year (**Heininger, 2006)**. The incidence of herpes zoster is very high in China, and it increases with age (**Macintyre, 2003)** and varies from country to country, ranging from 2.0 to 4.6/1000 person/year, without obvious geographic trend (Samuel, 2014; Pinchinat, 2013). VZV is highly infectious, and usually transmitted by droplets in the air and mucus in contact with the virus. Chicken pox is more common in children, while herpes zoster is more common in adults. Advanced age and dysfunctional cell-mediated immune responses are two well-established risk factors for VZV reactivation(**Thomas, 2004)**. Complications, however, do occur, particularly in infants, pregnant women and in immunocompromised individuals, including with postherpetic neuralgia (PHN)(Arnou, 2011). The incidence of VZV is not related to gender, but is closely related to season and climate, Spring and autumn are high-risk seasons. The incidence of VZV in tropical climate is low, but the average age of VZV infection is higher (**S al leras, 2000; Takahashi, 1992]**. Europe, America, Japan, Korea and China have adopted immunization strategies to control the epidemic of VZV through vaccination. In the sero-epidemiological investigation of VZV, Enzyme-linked immunosorbent assay (ELISA) is usually used to detect the concentration of varicella-zoster virus antibody (VZV-IgG) in serum. The concentration of VZV-IgG (> 100 mIU/mL) is positive, which indicates that the subjects have the ability to resist varicella virus infection.

With the immunity of the elderly decreasing, they are more susceptible to virus infection, so that VZV incidence increases and VZV would bring serious economic and spiritual burden to society. Because of high VZV incidence, VZV is included in the Chinese infectious diseases report. But the prevalence of VZV is not clear to people. The purpose of this study is to investigate the sero-prevalence of VZV in general population, analyze the influence factor of VZV infection, and provide theoretical basis for the prevention and treatment of VZV.

## 1. METHODS AND DATA

### 1.1 Search strategy

Literatures about the immune epidemic of varicella-zoster were searched on PubMed, the China National Knowledge Infrastructure (CNKI) database, WANFANG database and the Chinese Scientific Journals Full-Text Database (CQVIP), which published in recent 30 years in China. Chinese and English search terms are “Varicella-zoster virus”, “varicella”, “Herpes zoster”, “Varicella immunity” and so on. The list of all eligible studies and reviews was manually scanned to identify additional studies for inclusion. During extraction of publications, we avoided subjective bias by omitting names of the authors, journals, year, and country.

### 1.2 Inclusion criteria

All studies which aimed on VZV seropositivity in the Chinese population were included. Of the total publications, duplicate and similar ones were identified and excluded. Since both English and Chinese publications were included, publications with same data but different languages were removed in the next step. The title and abstract of the retrieved studies were independently screened by two investigators and the full texts of the selected studies were further evaluated according to the eligibility criteria. Any disagreements were resolved by consensus after consultation with a third reviewer. Inclusion criteria: (1) the Chinese and English literatures of study on varicella-zoster virus immunization epidemics in different regions of China; (2) The literature has obvious age stratification; (3) The subjects are general people; (4) VZV sero-prevalence in the population investigated by the literature research institute. Exclusion criteria: Any study that did not contain these information was excluded.

### 1.3 Data Extraction and Processing

Two evaluators independently retrieved documents and extracted data. Information about the study design, eligible population, age and gender distribution, inclusion and exclusion criteria were checked. When they disagreed, they could solve the problem by discussion or by soliciting opinions from third parties. Firstly, the data of all literatures were extracted and the age stratification interval was determined. The data was pooled according to age stratification (1-3, 4-6, 7-13, 14-19, 20-35, 36-45, > 45) by Microsoft Excel spreadsheet. The total prevalence, 95% CI confidence interval (Lower-Upper CL) and estimate were analyzed and calculated by open-meta-analysis software. Forest plots were drawn by Graphpad Prism, open-meta-analysis and RevMan 5.3 software.

### 1.4 Statistical Method

RevMan 5.3, Graphpad Prism and open-meta-analysis software was used for meta-analysis. The heterogeneity of the included studies was tested by χ^2^ test and Cochrane Q test. The heterogeneity was determined by *I*^*2*^ value, *I*^*2*^=0 was considered indicative of the variation between studies that was only caused by sampling error. *I*^*2*^<0.5 was considered indicative of a lack of significant heterogeneity among the included studies and a fixed-effects model was used for analysis. *I*^*2*^>0.5 was considered indicative of substantial heterogeneity and a random-effects model was used for analysis. Meta-analysis results are represented by forest plots.

Cochrane Q test was also used to test the heterogeneity of statistical results, and the significant level was at < 0.1. Heterogeneity calculation is the weighted sum of the square difference between the prevalence rate in individual studies and the combined prevalence rate in all studies, as well as the weights used in the aggregation method. We assume that the prevalence in all studies is the same as the invalid hypothesis. To test the heterogeneity, we calculated Q and compared it with the standard threshold table. (Allami, 2014).

The Ethics Committee of the First Afliated Hospital of Xian Medical University approved the study.

### 2. Results

### 2.1 Literature Retrieval Results

By searching Pubmed, CNKI, WANFANG and CQVIP databases, 2035 papers were obtained. By eliminating duplicate papers, 63 papers were preliminarily obtained through reading topics and abstracts, Which were in CNKI (n=21), Pubmed (n=39) and other methods (n=3). 13 incomplete literatures and 35 papers without age stratification were excluded by acquiring and reading the full text. Compare with the age stratification criteria, 5 literatures were excluded. According to the inclusion and exclusion criteria, 10 literatures (Cao, 1998; Wang, 2004; Li, 2016; Pan, 2003; Dong, 2017; Wu, 2015; He, 2011; Zhou, 2006; Zheng, 2000; Guo, 2007) were included in the meta-analysis. 11666 individuals in China were included, which was all in Chinese. A flowchart depicting the screening process of references at each stage is shown in Figure 1.

**Figure 1.**
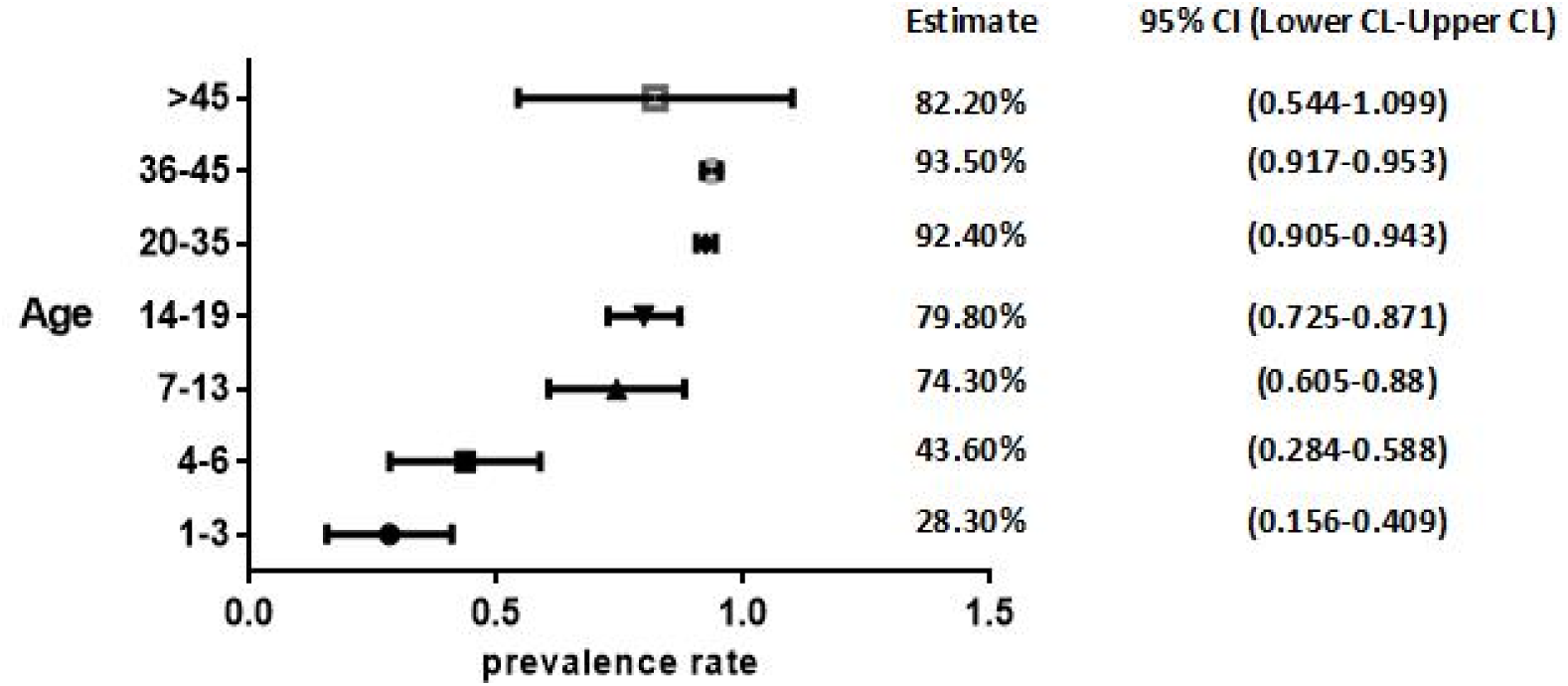
Varicella Immunization Forest Map at Different Age Levels (Prevalence Estimate)

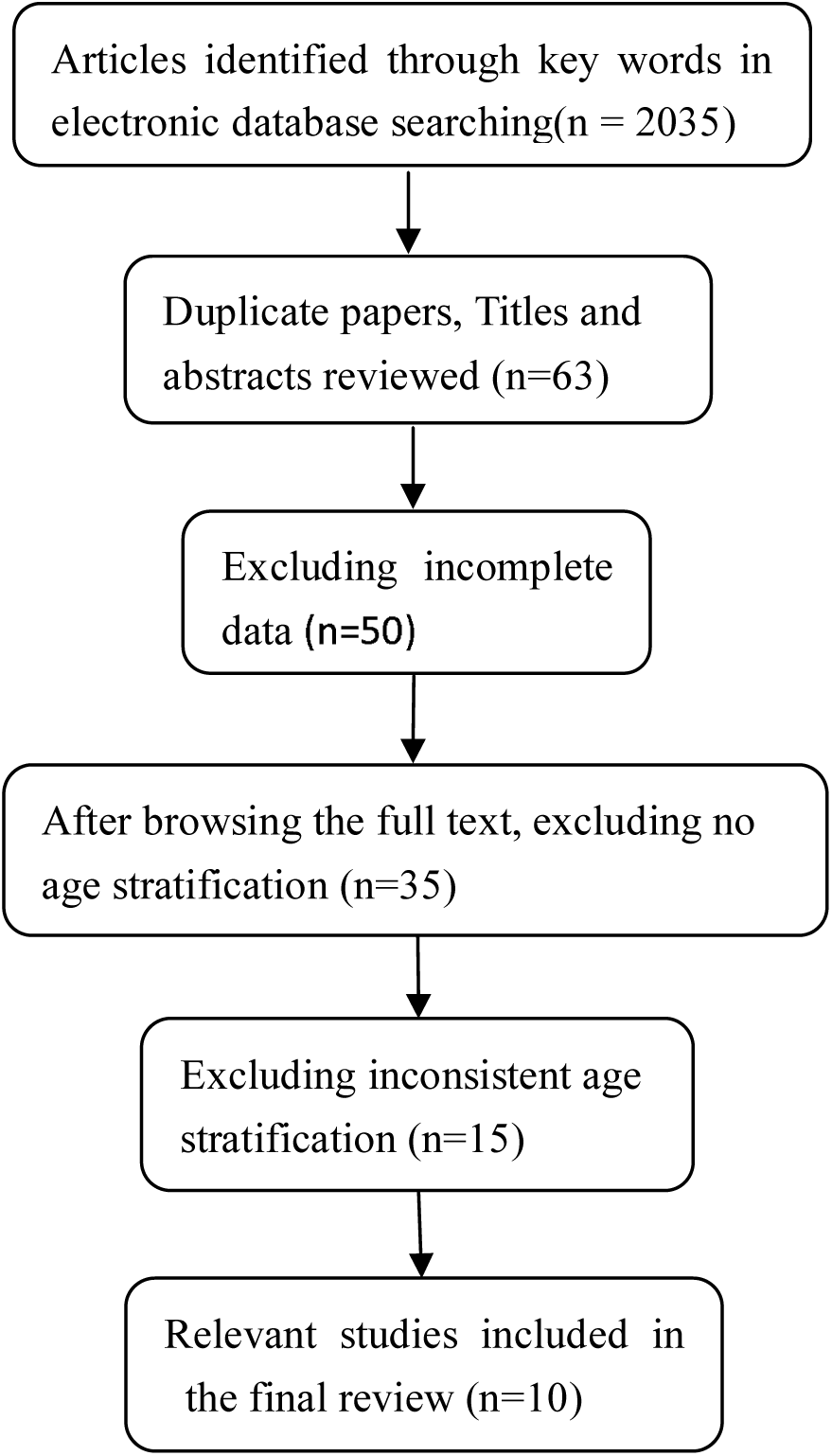

### 2.2 Basic information of included study

The level of VZV antibody in healthy Chinese population was studied in 10 papers, which research data is from 1997 to 2019. The subjects were surveyed from 10 provinces in the Chinese different geographical region (Shanghai, Guangdong, Beijing, Liaoning, Sichuan, Heilongjiang, Fujian, Zhejiang, Henan and Jiangsu), involving healthy children, residents, communities and rural populations. 11 666 individuals was included in this cross-sectional study. The sample size of each study ranged from 320 to 3073. There was no gender difference and the age of subjects was between 1 and 70 years old. Enzyme-linked immunosorbent assay (ELlSA) was used to detect anti-VZV-IgG antibodies in serum using ELlSA kit produced by BEEHRING, Germany. The basic information of included study was shown in Table 1.

**Table 1.**
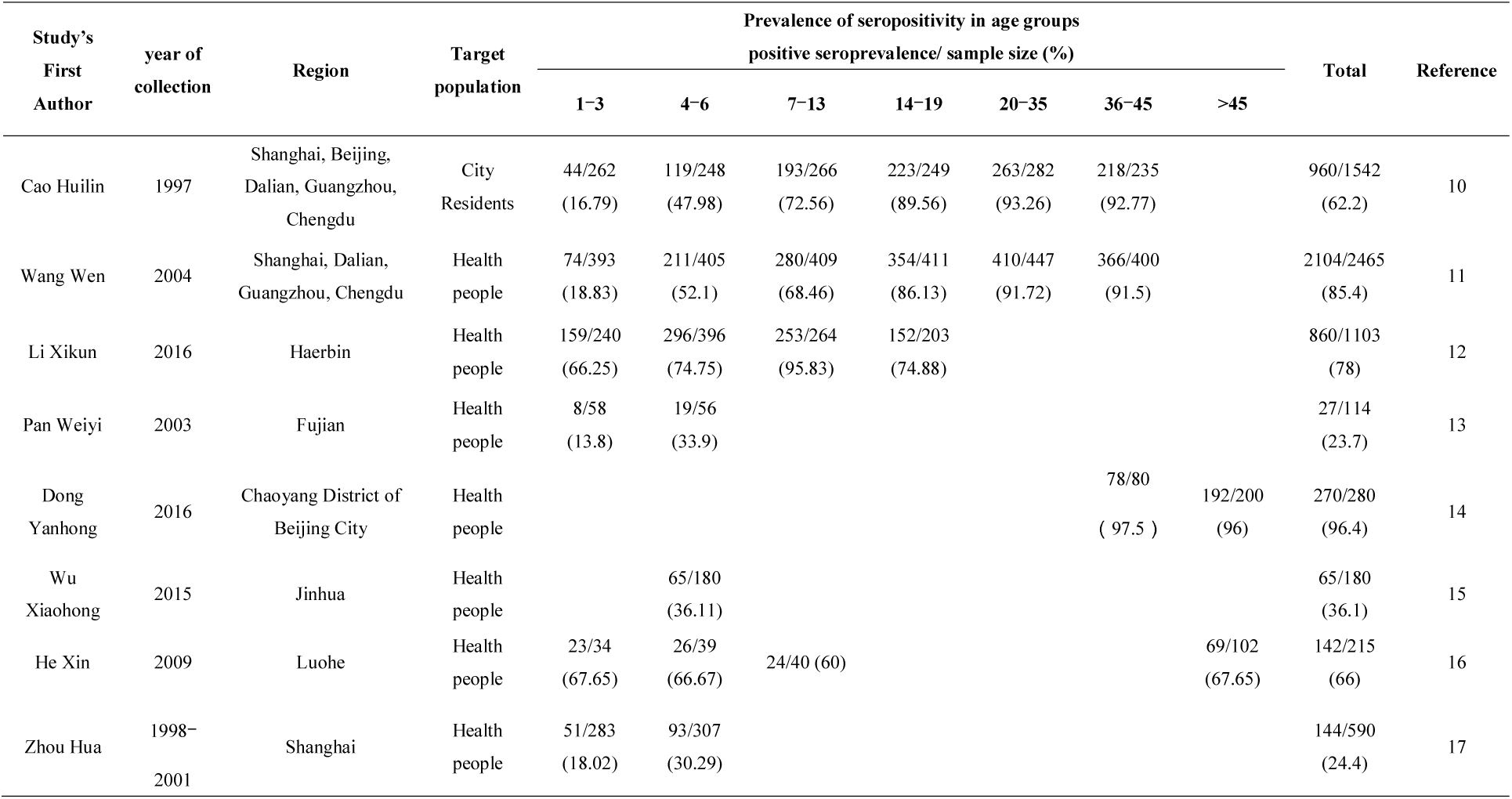
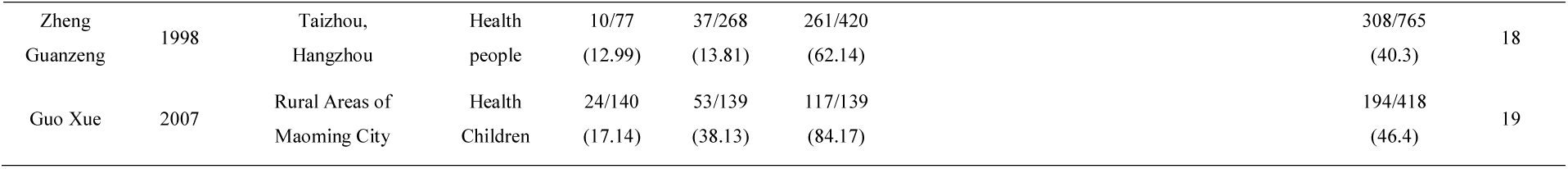
Basic information for inclusion in the study.

### 2.3 Analysis of Heterogeneity

10 studies were pooled and statistically analyzed with the age stratification, and the Q, *I*^*2*^, Q/df and P values of different age groups were calculated by random effect model and fixed effect model respectively. According to the statistical analysis method of heterogeneity, random effect model was used to do meta-analysis in the case of significant heterogeneity. The total prevalence and its 95% confidence interval were calculated by weighted average of individuals in 7 age groups (1-3, 4-6, 7-13, 14-19, 20-35, 36-45, and > 45 years old) (Table 2).

**Table 2.** Heterogeneity Analysis of 10 studies in Different Age Stratification.

| Age group | Random effect model |  |  |  | Heterogeneity | Fixed effect model |  |  |  |
| --- | --- | --- | --- | --- | --- | --- | --- | --- | --- |
| | Q | $I^2(\%)$ | Q/df | P | | Q | $I^2(\%)$ | Q/df | P |
| 1-3 | 262.342 | 97 | 37.477 | <0.001 | substantial | 262.342 | 97 | 37.477 | <0.001 |
| 4-6 | 459.584 | 98 | 57.448 | <0.001 | substantial | 459.584 | 98 | 57.448 | <0.001 |
| 7-13 | 250.033 | 98 | 50.007 | <0.001 | substantial | 250.033 | 98 | 50.007 | <0.001 |
| 14-19 | 82.3 | 98 | 41.15 | <0.001 | substantial | 82.3 | 98 | 41.15 | <0.001 |
| 20-35 | 0.604 | 0 | 0.604 | 0.437 | non-significant | 0.604 | 0 | 0.604 | 0.437 |
| 36-45 | 7.497 | 73 | 3.75 | 0.024 | substantial | 7.497 | 0 | 3.75 | 0.024 |
| >45 | 34.389 | 97 | 34.389 | <0.001 | substantial | 34.389 | 97 | 34.389 | <0.001 |

df=degrees of freedom. \*: significant P-value $\leq 0.10$ .

### 2.4 Prevalence of VZV

Data were collected from 10 studies according to age stratification (1-3, 4-6, 7-13, 14-19, 20-35, 36-45, and > 45) and gender. Prevalence estimates and 95% CI confidence intervals (Lower CL-Upper CL) were calculated using open-meta-analysis software (Table 3). Confidence areas were also calculated in Graphpad Prism software. The meta-analysis of point estimates and 95% confidence interval for VZV prevalence in different age groups were shown as a forest plot in Fig 1. The research results displayed that at the age of 1–3, 28.30% (95% CI; 0.156-0.409) of children showed to be infected. The seropositivity rate of VZV-IgG antibody sharply increased from 43.6% (95% CI: 0.284-0.588) at the age of 4-6 to 74.3% (95% CI: 0.605-0.88) at the age of 7-13. The peak prevalence was seen in the age 36-45 93.50% (95% CI; 0.917-0.953) while the VZV seroprevalence rate (82.20%, 95% CI: 0.544-1.099) was not increased in individuals of 45 and older. Trend of age-stratifed VZV seroprevalence rates in Chinese population during 1997 to 2019 is shown in Fig 1.

**Table 3.** Prevalence Estimates and 95% CI Confidence Intervals in Different Age Groups.

| Age group | Estimate | 95% CI<br>(lower CI-upper CI) | N of studies | Total sample |
| --- | --- | --- | --- | --- |
| 1-3 | 28.30% | (0.156-0.409) | 8 | 1487 |
| 4-6 | 43.60% | (0.284-0.588) | 9 | 2038 |
| 7-13 | 74.30% | (0.605-0.88) | 6 | 1538 |
| 14-19 | 79.80% | (0.725-0.871) | 3 | 863 |
| 20-35 | 92.40% | (0.905-0.943) | 2 | 729 |
| 36-45 | 93.50% | (0.917-0.953) | 3 | 715 |
| >45 | 82.20% | (0.544-1.099) | 2 | 302 |

Ten studies explored the seropositivity rate of VZV antibody, the estimates value of VZV prevalence and 95% confidence interval in each study were calculated and pooled (Fig 2). Due to substantial heterogeneity, random effect model was performed in the meta-analysis. The overall VZV seroprevalence in the Chinese population was 65.80% (95% CI; 56.5% - 75.1%). It is shown that 7616 individuals out of 11666 were seropositive of anti-VZV-IgG antibody(Fig 3).

**Figure 2.**
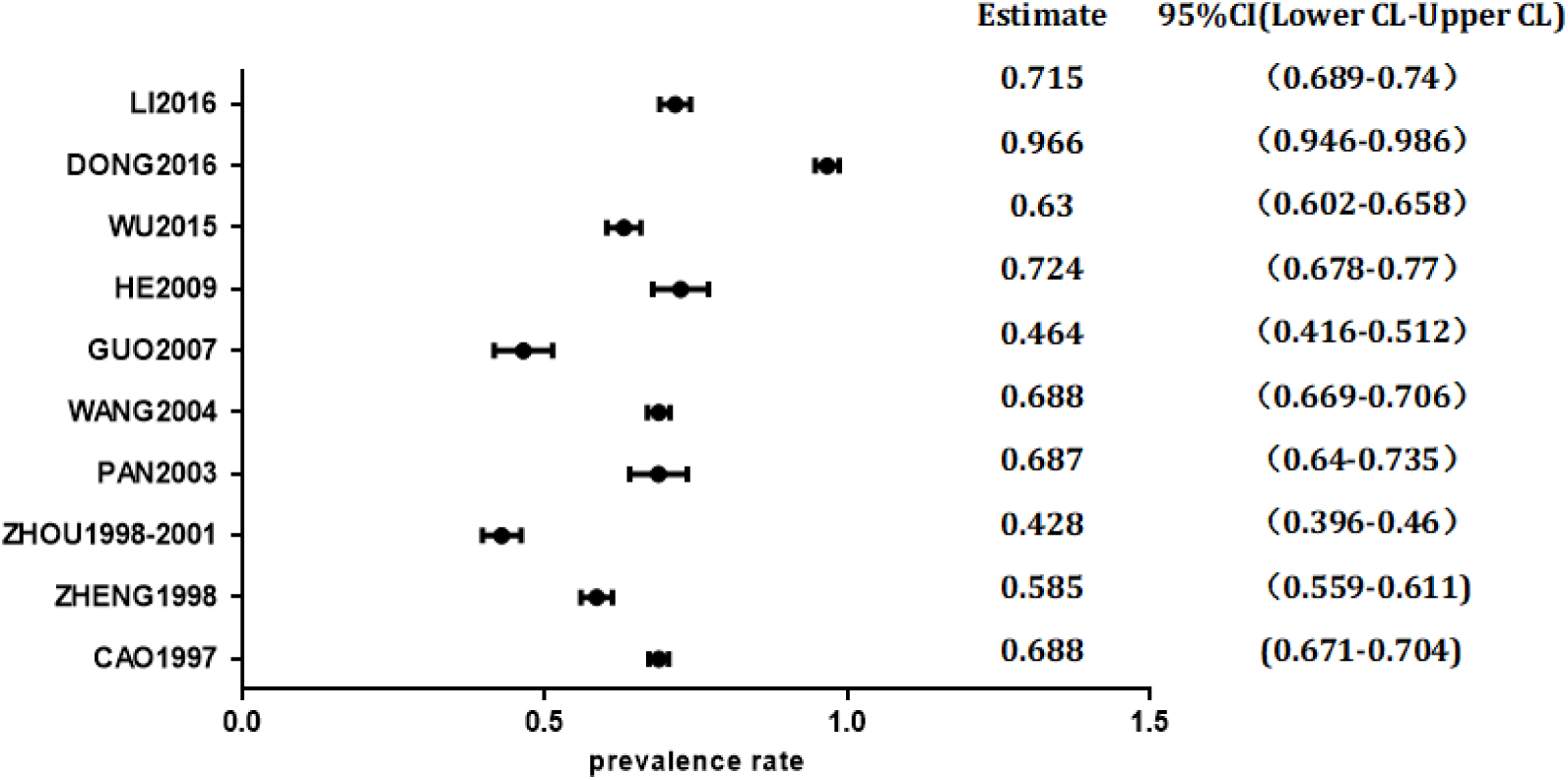
Forest charts of VZV prevalence in each study.

**Figure 3.**
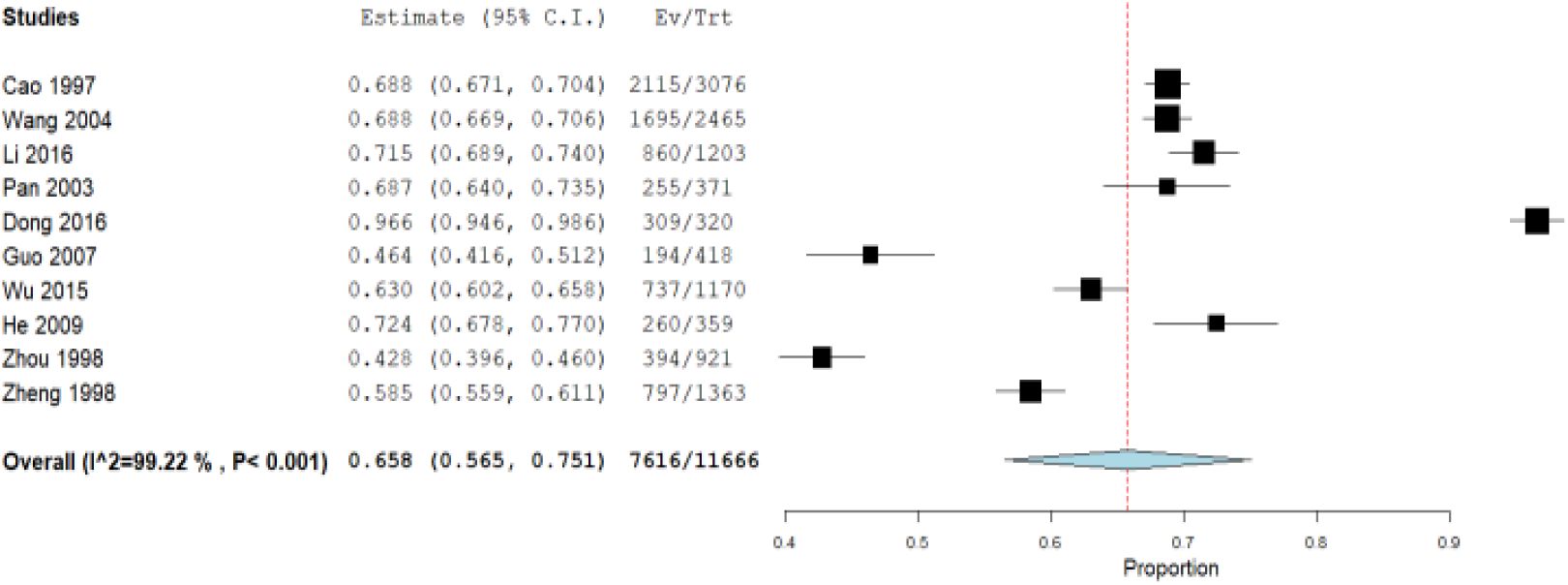
Forest plots of VZV prevalence in each study.

Nine studies observed the prevalence of anti-VZV antibodies in different sexes, which data was analyzed using Revman 5.3 software for meta-analysis. The results suggested that the prevalence of VZV in women was slightly higher than that in men, with no statistical significance (P>0.05) (Figure 4).

**Figure 4.**
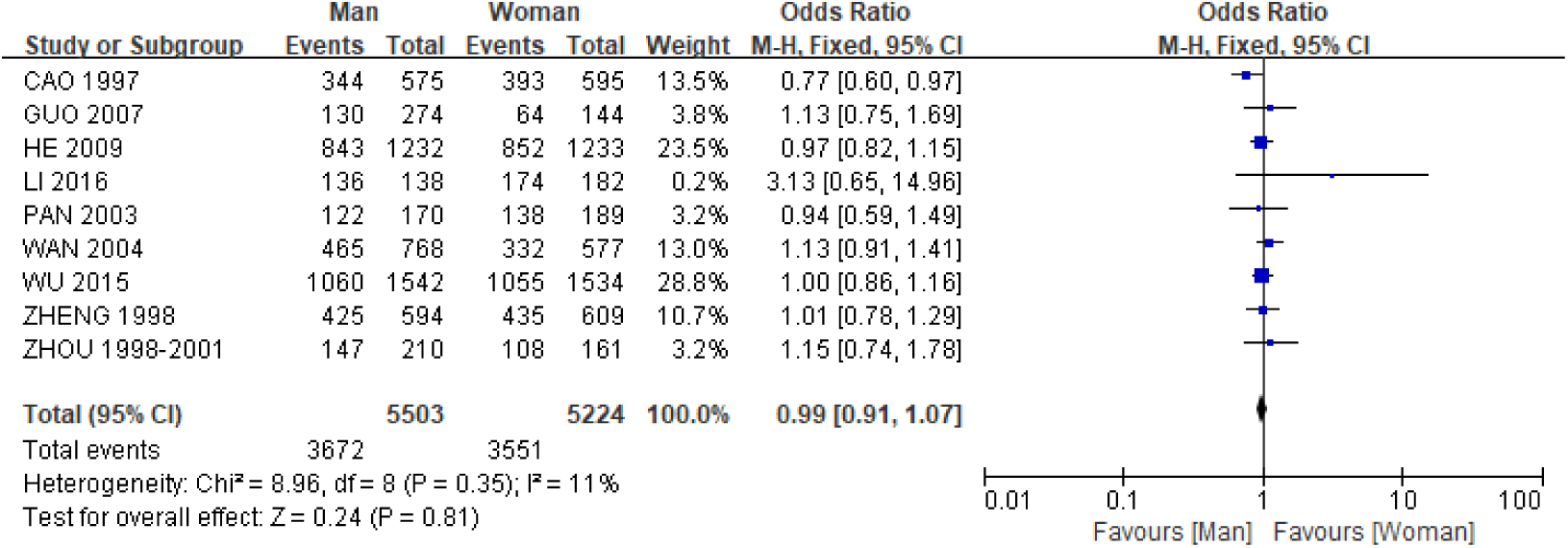
Forest Plots of VZV Prevalence in Different Genders.

## 3. Discussion

Varicella or herpes zoster is an acute infectious disease infected with VZV, and its incidence is very high all over the world. Human beings are generally susceptible to VZV. 90% of the susceptible people who come into close contact with VZV are infected mainly through respiratory tract. Immunity occurs after illness, and no secondary infection appeared. In vivo antibodies can not eliminate latent varicella virus in the spinal nerve root ganglia, and herpes zoster can still occur many years later(**Liu, 2010)**. The predilection age of varicella is infants and children, while herpes zoster is a recurrent disease, usually seen in adults aged 40 to 70. There are more cases in spring and autumn.

10 studies on VZV immunity prevalence were included in this meta-analysis. Open-meta-analysis software was used to collect and analyze the results according to age stratification. Cochrane Q test and *I*^*2*^ test were used to evaluate the heterogeneity of the results. The results showed that the prevalence of VZV in all age stratification had greater or lesser heterogeneity. In all reports, an age-dependent pattern was demonstrated in VZV seropositivity. Seropositivity rates were low in the early childhood as the frequency of positive samples was 28.30% (95% CI; 15.6-40.9) during the first 1-3 years. The VZV seroprevalence increased from 28.3% in 1-3 years old to 43.6% in 4-6 years old. The VZV prevalence at the age of 7-13 years old was similar to that at the age of 14-19 years old, 74.3% and 79.8% respectively, and was about three times higher than that of 1-3 years old and more than 30% higher than that of 4-6 years old. By the age 20-35 years, VZV seroprevalence of young people steeply rose to 92.4% and at the age of 36-45 years, 93.5% of the population had already been infected by VZV. VZV prevalence of 36-45 years old was close to that of 20-35 years old. Finally, 82.2% of elderly people were seropositive for anti-VZV antibodies by the age of 45 and more, which was lower than that of the age group of 20-45 years old. There was some heterogeneity in the VZV prevalence of 1-3, 4-6, 7-13, 14-19, 36-45 and over 45 years old. Random effect model was used to analyze the data and there was significant statistical significance. No heterogeneity in the statistical results of VZV prevalence is between 20 and 35 years old, and fixed effect model (*I*^*2*^ = 0, *P* = 0.437) was used to meta-analysis. The results of this study was consistent with those of Allami et al. (Abbas, 2014), but the age stratification of the two studies is different. The prevalence of VZV in Iran is 89.54% in the age group of 26-30 years old, followed by 85.92% in the age group of 31-40 years old. The VZV prevalence of children in China is 28.3% in the age group of 1-3 years old, 43.6% in 4-6 years old, and 74.3% in 7-13 years old, which is higher than Iran (21.96% in 1-5 years old, 42.09% in 6-10 years old, 59.44% in 11-15 years old). The prevalence of VZV in both studies over 40 years old is over 80%. It is found that the seroprevalence of VZV increased with age before the age of 45, which is approved by results of Mahamud et al. (Mahamud, 2014). Before introducing the routine varicella vaccination program, the VZV seroprevalence among students of American Samoa University and elementary school increased with age, from 76.0% at the age of 4-6 to 97.7% by the age of 23 and more. In contrast to the research results of Pinchinat et al. (Pinchinat, 2013), Pinchinat found that the incidence of herpes zoster in Europe increased with age and became more serious after age 50.

Ten studies were distributed in six southern provinces and cities (Zhejiang, Shanghai, Fujian, Guangdong, Jiangsu, Sichuan) and four northern provinces and cities (Beijing, Liaoning, Heilongjiang, Henan). According to the VZV prevalence in each study, open-meta-analysis software was used to conduct a summary analysis, and the heterogeneity of each study was evaluated by Cochrane Q test and *I*^*2*^ test. Eight studies were selected to independently study the prevalence rate of VZV in a certain region. The results found that compared with that in the southern region, the VZV prevalence in the northern region (Harbin 71.5%, Beijing Chaoyang 96.6% and Luohe 72.4%) was higher than that in the southern region (Fuzhou 68.7%, Jinhua 63.0%, Shanghai 42.8%, Hangzhou and Taizhou 58.5%, and Maoming 46.4%), which was basically similar to the VZV seropositive rate between tropical and temperate regions in Iraq(Allami, 2014).

By analyzing the influence of gender on the VZV prevalence in nine studies, it was found that the average prevalence of VZV in women was 62.71%, and that in men was 62.07%. The VZV prevalence in women was slightly higher than that in men (*P*>0.05, no statistical difference), which was consistent with the results of Pinchinat et al. (Pinchinat, 2013). The incidence of VZV in women was also higher than that in men, and the difference increased with age. Compared with Amjadi et al. (Amjadi, 2017), the prevalence rate of VZV in Chinese women was 62.71%, which was lower than that in Iranian women (80.47%). Dong Yanhong et al. (**Dong, 2017)** found that the VZV prevalence of the elderly that was 96.56% in Chaoyang District of Beijing was slightly higher than that of 95.30% in Iran. Due to the decrease of immunity, the elderly was more susceptible to VZV infection.

This Chinese study with total VZV seropositivity rate 65.8% (95% CI:56.5-75.1%) came to a same conclusion with Iranian’s (78.50%) and Polish(76.6%) rate of VZV prevalence (Amjadi, 2017; Siennicka *et al*.,2009).

## 4. Conclusion

In summary, according to the inclusion and exclusion criteria, this study included 10 studies on the prevalence of VZV published from 1997 to 2017 in different regions of China. Results from this study demonstrated a close relationship between seroprevalence and age, but no close correlation with gender, rural/urban areas and geographical regions of China(Siennicka *et al*., 2009). The meta-analysis showed that the VZV seroprevalence increased with age, and there was no significant difference between different genders. The prevalence of VZV in northern areas was higher than that in southern areas. From 1997 to 2019, there was a tendency that the VZV prevalence was increasing year by year, so the public health prevention of VZV needs to be further strengthened to benefit people’s health. The seroepidemiological survey of VZV is helpful to understand the status of VZV infection in healthy people and to assess the risk of herpes zoster.

## Data Availability

none

## Author Contributions

Wang L. and Zhang M. X. and Li Z. W.: manuscript writing and revising and final approval of the manuscript; Shao X. L. and Sun F.: pooling and analyzing data and final approval of the manuscript; Zhang H. Y., Liu S. Y. and Lu B.: searching and extracting data and final approval of the manuscript; All authors have read and approved the final manuscript.

## Acknowledgements

This study was supported in part by grants from Key Research & Development Program Projects of Shaanxi Province, China (2015SF080, 2017SF-336, 2019SF-163) and The Medical Project of Xi’an Science and Technology Bureau, China(2017122SF/YX016 (5)).

## Ethical approval

This study is exempt from ethical approval because it will be collecting and synthesizing data from previous reports in which informed consent has already been obtained by their investigators.

## references

1. Davison AJ, Scott JE. The complete DNA sequence of varicella-zoster virus. J Gen Virol 1986; 67:1759–1816. doi:10.1099/0022-1317-67-9-1759

2. Heininger U, Seward JF. Varicella. Lancet 2006; 368:1365–76. doi:10.1016/S0140-6736(06)69561-5

3. Macintyre CR,Chu CP, Burgess MA. Use of hospitalization and pharmaceutical prescribing data to compare the prevaccination burden of varicella and herpes zoster in Australia. Epidemiol Infect 2003; 131: 675–682. doi: 10.1017/s0950268803008690

4. Samuel LK. Varicella and herpes zoster vaccines: WHO position paper, June 2014. Wkly Epidemiol Rec 2014; 89:265–287. PMID: 24983077

5. Pinchinat S, Cebrián-Cuenca AM, Bricout H, et al. Similar herpes zoster incidence across Europe: results from a systematic literature review. BMC Infectious Diseases 2013; 13:170. doi: 10.1186/1471-2334-13-170

6. Thomas SL, Hall AJ. What does epidemiology tell us about risk factors for herpes zoster? Lancet Infect Dis 2004; 4, 26–33. doi: 10.1016/S1473-3099(03)00857-0

7. Arnou R, Fiquet A, Thomas S, et al. Immunogenicity and safety of ZOSTAVAX□approaching expiry potency in individuals aged ≥50years. Hum Vaccin 2011; 7:1060–1065. doi:10.4161/hv.7.10.16480

8. S al leras L Dominguez, Vidal J. Sero-epidemiology of varicella-zoster virus infection in C at alonia (Spain): Rationale for universal vaccination programmer. Vaccine 2000; 19:183–188. doi: 10.1016/S0264-410X(00)00178-X

9. Takahashi M. Current status and prospects of live varicella. vaccine 1992; 10:1007–1014. doi: 10.1016/0264-410X(92)90109-W

10. Allami A, & Mohammadi N. Varicella immunity in Iran: an age-stratifed systematic review and meta-analysis. Iran J Microbiol 2014; 6:372–381. PMID: 25926953

11. Cao HL D. J, Jia ZY, et al. The prevalence of varicella-zoster virus in some areas of China. Chinese Journal of Vaccines and Immunization 1998; 1:42–44.

12. Wang W, Liu D, Li DJ. Survey on the positive rate of varicella-zoster virus antibody in four urban populations. Journal of Preventive Medicine Information 2004; 2:154–155. doi: 10.3969/j.issn.1006-4028.2004.02.022

13. Li XK, Gao XL, Li Y, et al. Seroepidemiological Survey of Varicella in Healthy Population Aged 1-19 in Harbin in 2016. Chinese Journal of Contemporary Pediatrics 2019; 3:203–207.

14. Pan WY, Zhang XD, Cai ZK, et al. Seroepidemiological survey of varicella-zoster virus in healthy population of Fujian Province. Practical preventive medicine 2003; 6:864–865. doi: 10.3969/j.issn.1006-3110.2003.06.012

15. Dong YH, Shi NM, Li LL, et al. Survey of varicella-zoster virus antibody level among healthy population in Chaoyang District of Beijing. Chinese Journal of Biologicals 2017; 5: 509–13.

16. Wu XH, Zhu SY, Pang ZF, et al. Survey of varicella-zoster virus antibody level in healthy population of Jinhua City in Zhejiang Province in 2015. Chinese Journal of Vaccines and Immunization 2016; 3:281–84. doi:

17. He X, Liu C, Li NX, et al. Seroepidemiological survey of varicella-zoster virus among healthy people in Luohe. Modern Preventive Medicine 2011; 4:601–602+605. doi:

18. Zhou H, Wang SQ, Chu Y, et al. Survey report on varicella-zoster virus infection rate among healthy population in some areas of Shanghai. Chinese Journal of Vaccines and Immunization 2006; 2:137–139. doi: 10.3969/j.issn.1006-916X.2006.02.019

19. Zheng GZ, Qiu DH, Yao PP, et al. Seroepidemiological study of varicella virus. Chinese Journal of Public Health 2000; 7:31–33. doi: 10.11847/zgggws2000-16-07-25

20. Guo X, Zheng HY, Liu L, et al. Survey of varicella-zoster virus infection and antibody level in rural areas of Guangdong Province. Chinese Journal of Health Laboratory Technology 2007; 2: 334–335. doi: 10.3969/j.issn.1004-8685.2007.02.062

21. Liu YN. Clinical Study on Different Acupuncture and Moxibustion Methods for Herpes Zoster [D]. Hubei University of Traditional Chinese Medicine, 2010. doi:10.7666/d.y1711878

22. Abbas A, Navid M. Varicella immunity in Iran: an age-stratifed systematic review and meta-analysis. Iran J. Microbiol 2014; 6:372–381. PMID:25926953

23. Mahamud A, Leung J, Masunu-Faleafaga Y, et al. Varicella zoster virus in American Samoa: seroprevalence and predictive value of varicelladisease history in elementary and college students. Epidemiol Infect 2014; 142:1002–1007. doi: 10.1017/S095026881300174X

24. Amjadi O, Rafiei A, Haghshenas M. A systematic review and meta-analysis of Sero-prevalence of Varicella Zoster virus: A nationwide population-based study. J Clin Virol 2017; 87:49–59. doi: 10.1016/j.jcv.2016.12.001

25. Siennicka J, Trzcińska A, Rosińska M, et al. Seroprevalence of varicella-zoster virus in Polish population. Przegl Epidemiol 2009; 63:495–499. PMID:20120946

